# From individual to population disease risk: population-averaged polygenic risk scores

**DOI:** 10.1101/2022.06.20.22276484

**Authors:** Simin He, Liuqing Peng, Jiarui Jing, Juping Wang, Tong Wang

## Abstract

Polygenic risk score (PRS) is a tool to quantify the susceptibility of an individual to a disease by using the results of individual gene sequencing to calculate the cumulative effect of genetic susceptibility loci. In this study, we extend the individual-based PRS to the population-based mean polygenic risk score (PMPRS), Further more, we constructed the risk score ratio (RSR) for etiology exploration. In the case study, we applied above method to explore the relationship between study six inflammatory factors and coronary heart disease (CHD). Our studies have shown that tumor necrosis factor-α (TNF-α) and procalcitonin (PCT) are potential risk factors for CHD. The RSR corresponding to TNF-α is 4.97 (prediction interval: 2.99, 10.548); the RSR corresponding to PCT is 21.87 (prediction interval: 2.29, 232.28), and controlling the levels of TNF-α and PCT can be used as a primary preventive measure to reduce the occurrence of coronary heart disease.

## 1. Introduction

The development of genetics has provided great convenience to human genetic research. With the continuous advancement of biotechnology, the economic cost of gene sequencing has decreased, making genetic research no longer an expensive and high threshold content. At the same time, the development of genetic technology has also provided multi-level and multi-dimensional medical big data for biomedical research. There are not only molecular features including genome level, epigenome level, transcriptome level, proteome level, metabolome level, etc., but also phenotypic characteristics at the organism level. Data covering all levels from genetic variation to characterization of living organisms provides a valuable resource for the study of complex diseases^[1]^. Genome-wide association studies (GWAS) identifies single nucleotide polymorphisms (SNPs) associated with complex diseases by examining differences in millions of SNPs across the genomes of diseased and healthy individuals. It provides new ideas and methods for revealing the genetic basis of complex diseases, identifying susceptibility functional regions, and exploring gene structures^[1]^.

Among the non-communicable diseases that have a major impact on public health, the genetic basis of most diseases is polygenic, including hundreds or thousands of genetic variants (or polymorphisms)^[2]^. Each genetic variant associated with a disease provides different information in indicating the gene or biological pathway associated with that disease. Therefore, we hope to use as comprehensive genetic information as possible to predict disease risk, so that it has wider clinical utility.Risk score is an important evaluation method for disease risk prediction in epidemiological research^[3]^. Among them, the genetic risk score (GRS) uses genetic variation to build a model and evaluates the effect of genetic susceptibility in the risk prediction model^[4]^. Since the occurrence of most complex diseases is controlled by multiple loci, the effect of a single or only a few loci on the disease is weak. In order to better assess the risk of disease, it is necessary to integrate the information of polygenic loci, that is, to perform a polygenic risk score (PRS)^[2]^.

PRS refers to an assessment tool that quantifies the susceptibility of an individual to a disease by calculating the cumulative effect of multiple susceptibility loci^[5]^. When an individual has a higher PRS on a disease, the greater the probability that he will have the disease in the future. Since the occurrence of most diseases is the result of the interaction of genetics and environment, only when the genetic risk and environmental risk are high at the same time, the individual will show the disease state. Therefore, although PRS represents an individual’s genetic prediction of disease, PRS is more used to prevent disease by changing adverse factors in the environment earlier.^[6]^. For example, it is found that a subject has a very high genetic risk score for type 2 diabetes through gene sequencing and calculation of PRS. In this case, the subject can prevent the occurrence of diabetes through early adjustment of diet and living habits^[7]^.

With the rapid development of biotechnology, PRS has gradually become a widely used tool. However, since the PRS is based on the genetic testing results of the subjects, the interpretation of the results is limited to the subjects themselves, which limits the application scope and application value of the research results to a large extent^[8]^. From the perspective of public health promotion, we constructed a population-based Mean Polygenic Risk Score (PM-PRS). Based on PRS, PM-PRS extends individual risk prediction to population risk prediction. Firstly, we calculate the risk allele expectation in the study population based on the minor allele frequency (MAF) of the population. Then, we estimate the average disease risk score for the study population based on the above expected value. PM-PRS extends disease risk estimation from individual level to the population level. We aim to promote early detection and intervention of high-risk and sensitive groups, and we hope to provide scientific guidance for “tertiary prevention” of populations. At the same time, it helps to strengthen in-depth research on the differentiation of diseases between different populations.On the basis of PM-PRS, we further constructed the risk score ratio (Risk Score Radio, RSR) to explore the potential etiology of the disease from the perspective of genetics. Finally, we applied the RSR to explores the role of the inflammatory hypothesis in the development of coronary heart disease (CHD) in order to provide guidance and help for the prevention of CHD from a genetic perspective.

## 2. Method

### 2.1 Polygenic risk score

Gene polymorphism refers to the phenomenon that the structure or nucleotide sequence of the same gene is not exactly the same among different individuals, which can be regarded as a high-frequency manifestation of allelic variation. Although gene polymorphism does not necessarily affect gene function, it can be used as a marker to distinguish individuals^[7]^. Alleles can be divided into mutant type and wild type according to the mutation of the gene locus. Among them, the wild type is the unmutated allele, also known as the major allele, and the mutant genotype is the mutated allele, also known as the minor allele. In the exploration of disease etiology, genetic mutation is often regarded as a risk factor for disease occurrence, so the mutated gene is also called risk allele.

PRS quantifies an individual’s susceptibility to disease by calculating the cumulative effect of multiple susceptibility loci^[5]^. It is the sum of the number of risk alleles which were weighted by the effect size of each allele on the disease estimated by GWAS^[9]^, and the formula is as follows:

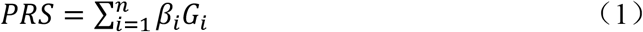

In the above formula, *i* represents the SNP, *n* represents the number of SNPs that have an impact on the disease, *β*_*i*_ represents the effect value of the *i*-th SNP on the disease, *G*_*i*_ and represents the expression number of the minor allele of the subject at the SNP locus (for biallelic polymorphism, the expression numbers of no mutation, heterozygous mutation, and homozygous mutation are 0, 1, and 2, respectively).

In the PSR, *β*_*i*_ is mostly provided by GWAS, and *G*_*i*_ is obtained from the individual genetic testing result. In order to avoid the bias caused by ethnic differences and sample overlap, it is necessary to ensure that the individual tested and the GWAS sample providing the effect value are from the same ethnicity but different populations^[1]^.

### 2.2 Population-based Mean Polygenic Risk Score

Minor Allele Frequency (MAF) is a concept in population genetics that refers to the frequency of a minor allele (the second most common genotype) in the population. MAF provides information for distinguishing common variants from specific variants in a population^[10]^. Borrowing the population genetic variation information contained in MAF and its ability to distinguish different groups, we construct the PM-PRS, which extends the concept and application of individual polygenic risk scores to population polygenic risk scores.

In the assessment of population disease risk, the association between SNPs and diseases can be easily obtained from GWAS. Therefore, in the assessment, the focus is on estimating the expression of risk genes in the population Since the vast majority of SNPs are biallelic polymorphisms (that is, alleles are only expressed in major alleles and minor alleles). As described above, on a specific SNP, the expression levels of no mutation, heterozygous mutation, and homozygous mutation are 0, 1, and 2, respectively. At this point, combined with MAF, we can calculate the probability of occurrence of three genotypes with different expression levels. which is:

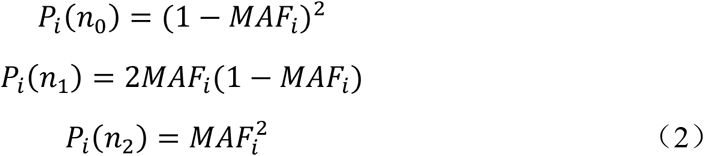

In the formula, *n*_0_, *n*_1_, *n*_2_ represent the number of alleles at the *i*-th SNP respectively.

According to the probabilities of the three different expression levels in (2), the expected value of the expression level of the minor allele at the *i-*th locus can be calculated as follows:

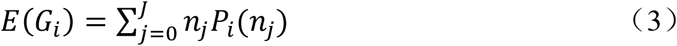

The minor allele expression is expected to represent the average expression of the minor allele at the *i*-th SNP locus in the sample population. When the expected value of the minor allele expression at the *i*-th SNP locus in the studied sample population is higer, it means that the population has a higher possibility of gene mutation at this locus.

From this, we replaced the in the individual PRS with the population minor allele expression expectation to obtain the PM-PRS. The formula is as follows:

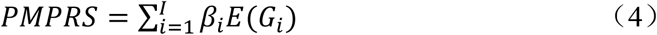

In the above formula, *β*_*i*_ is the same as in PRS, that representing the effect value of the *i*-th SNP on the disease. The PMPRS reflects the disease risk of the sample population. The higher the score, the higher the risk of the disease in the sample population. In addition, according to the linear derivation of variance, the variance of PM-PRS is as follows:

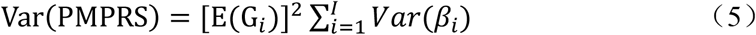

### 2.3 Risk score ratio

Exploration of etiology plays a crucial role in the study of the formation mechanism, prevention, control and treatment of the disease. Limited by financial and ethical constraints, randomized controlled trials (RCTs), which regarded as the “gold standard” for etiological inference, have not been fully and widely used in actual etiological inference. On the other hand, although observational research based on the real world has the advantage of saving time and effort, the reliability of the results cannot be fully guaranteed due to the control of confounding factors or the problem of reverse causality. Benefiting from the characteristic that genes are congenitally formed, they are not easily affected by confounding factors and there is almost no reverse causal when used for etiology exploration. Taking advantage of above advantage of genetic variation, we propose the RSR and apply it to the exploration of potential risk factors for disease. First, we calculated the risk score for the exposure and diseases by the genetic variation they shared. Second, we calculated ratios based on the risk scores between exposures and outcomes. The formula is as follows:

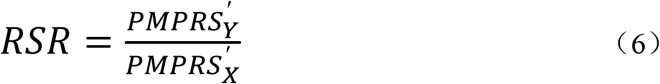

The RSR can be understood as the point of the population that increases the risk of disease when a genetic factor shared between the exposure and the outcome increases the risk of exposure by 1 point in the sample population.When the RSR>1, the exposure factor may be a potential risk factor for the disease; and when the RSR<1, it indicates that the exposure factor may be a potential protective factor for the disease.

In the estimation of RSR, we make the following assumptions:

1) 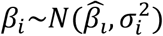, where 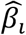 is the true causal effect between the SNP and the trait; 2) *MAF*∼*U* (0,0.5). According to the above two assumptions, and since PMPRS is a linear combination of *β*_*i*_ and MAF, PMPRS also follows a normal distribution, that is,. In addition, the RSR is obtained by the ratio of the risk score of disease to exposure, so the RSR follows the Cauchy distribution, that is, 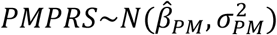 *RSR*∼*C*(*γ,χ*_0_). Since the non-convergence characteristics of the Cauchy distribution, the RSR has no expectation and variance, and its prediction interval cannot be obtained. In this study, we used the Bootstrap to estimate the prediction interval for RSR by resampling 1000 times.

## 3. Case Study: Exploring the Potential Causes of Coronary Heart Disease

Coronary heart disease (CHD) is a kind of chronic disease that threatens human life and health. According to the data published in the “China Health Statistical Yearbook” released in 2020, the CHD mortality rates of urban and rural residents in China in 2019 were 1.2159/100,000 and 1.30.14/100,000 respectively^[11]^. In recent years, with the accelerating pace of work and life, the prevalence of CHD has also increased year by year. CHD and its complications such as heart failure and embolism also bring huge health and economic burdens to the patient’s family and society.

Inflammation is the body’s defense response to external stimuli. When the body is attacked by infection or non-infectious factors, the inflammatory mechanism responds quickly and protects our body. However, when the body’s defense response is overdone, a response that damages the body’s own tissues may occur^[12]^. The inflammatory hypothesis is an important hypothesis for the exploration of the etiology of CHD. The inflammatory hypothesis states that the occurrence of CHD is first initiated by dysfunction or damage of vascular endothelial cells. At this time, the body’s defense mechanism responds rapidly and induces a large number of inflammatory cells to enter and stay in endothelial cells^[13]^. Apolipoprotein B-rich lipoproteins (low-density lipoprotein, very low-density lipoprotein remnants, chylomicron remnants, etc.) also enter and attach to damaged endothelial cells, leading to the formation of atherosclerotic plaques and progress^[14]^. At the same time, some studies have found that with the continuous development of inflammation, the vascular plaque is gradually unstable. The occurrence of plaque rupture, fissure or erosion will further induce thrombosis and lead to myocardial injury and necrosis, which means that inflammation is involved in the whole process of CHD^[15]^. In addition, recent studies have shown that immune cells dominate early atherosclerotic lesions, their effector molecules accelerate the progression of lesions, and the activation of inflammation can cause acute coronary syndrome^[16]^.

In recent years, the exploration of the correlation and causal association between inflammatory factors and CHD has emerged in an endless stream. In the study of the association between C-reactive protein (CPR) and fibrinogen and CHD, the results of a case-control study support the conclusion that CRP is an inflammatory marker of activation of CHD^[17]^. However, human genetic evidence has denied the possibility of a causal relationship between these liver-derived inflammatory factors and CHD^[18]^. In contrast, among “upstream” inflammatory markers (eg, pro-inflammatory cytokines, etc.), there may be a more direct etiological relationship with CHD due to their direct control of the inflammatory cascade^[19]^. Among the “upstream” inflammatory markers, interleukin-6 (IL-6) has been widely demonstrated to be associated with CHD^[20-22]^, and genetic evidence also supports the role of IL-6 in CHD. causal role in^[23]^. At the same time, the association between other “upstream” inflammatory markers and CHD has gradually attracted widespread attention. Existing studies have confirmed that tumor necrosis factor-α (TNF-α), as an important adipo-inflammatory factor, plays an important role in the occurrence and development of CHD and atherosclerotic diseases^[24, 25]^; Esther Lutgens et al. found that in the treatment of CHD and atherosclerosis, targeted therapy of interleukin-1β (IL-1β) can prevent the occurrence of cardiovascular events^[26]^.

The inflammatory hypothesis of CHD has been proposed for nearly 20 years, however, the causal association between CHD and related inflammatory markers has not yet reached a consistent conclusion. In order to explore the inflammatory hypothesis in CHD, this study investigated the causal associations between “upstream” inflammatory markers and “downstream” inflammatory markers and CHD. We hope to provide a scientific basis for the causal association of the inflammatory hypothesis in CHD from a genetic perspective.

### 3.1 Data Sources

The potential etiologies included in this study were inflammatory markers, including “upstream” inflammatory markers (IL-1β, IL-6 and TNF-α) and “downstream” biomarkers (CRP, procalcitonin (PCT) and Serum amyloid A (SAA)). The gene sequencing summary data used to provide the correlation coefficient between genetic variation and traits in this study were all from the latest published GWAS, and the specific information is shown in Table 1. The MAF information was obtained from the UK10K study, which is based on the Avon Longitudinal Study of Parents and Children (ALSPAC). The cohort collected baseline information on more than 14,000 mothers during pregnancy since 1991-1992, and conducted a multi-year follow-up survey of parents and children in the cohort, including a large amount of genetic and environmental information. The MAF information applied in this study was derived from the sequencing results of the whole-genome cohort in UK10K^[27]^.

**Table 1.** Sources of research data

| Traits | Sources | Sample size | PMID |
| --- | --- | --- | --- |
| CHD | UKBiobank | 361194 | - |
| IL-1β | INTERVAL study | 3301 | 29875488<sup>[28]</sup> |
| IL-6 | INTERVAL study | 3301 | 29875488<sup>[28]</sup> |
| TNF-α | INTERVAL study | 3301 | 29875488<sup>[28]</sup> |
| CRP | KORA study | 1000 | 28240269<sup>[29]</sup> |
| PCT | INTERVAL study | 3301 | 29875488<sup>[28]</sup> |
| SAA | INTERVAL study | 3301 | 29875488<sup>[28]</sup> |

Note: CHD: coronary heart disease; IL-1β : interleukin-1β ; IL-6: interleukin-6; TNF-α : tumor necrosis factor-α ; CRP: C-reactive protein; PCT: procalcitonin; SAA: serum amyloid A.

### 3.2 Statistical methods

The study screened the included SNPs by *p* < 5e-8, and we explored potential associations between the six studied inflammatory markers and CHD according to the included SNPs. Statistical analysis was performed in this study using R 4.0.5 software. We also generated the PMPRS package (https://github.com/PLQ74/PMPRS) for application.

### 3.3 Results

Table 2 shows the RSR results of six inflammatory factors to CHD. The results shows that IL-6 and TNF-α may be potential risk factors for CHD among the “upstream” inflammatory markers. While according to the prediction interval results, only TNF-α was identified as a risk factor for CHD. In the European sample population included in the study, when the shared genetic factors between TNF-α and CHD caused a 1-point increase in the risk of TNF-α, they increase in the risk of CHD in the population is 4.97. Among the “downstream” inflammatory markers, the results showed that PCT may be a potential risk factor for CHD. When the shared genetic factors between PCT and CHD increased the risk of PCT by 1 point, the risk of CHD increased by 21.87 points. We also draw a forest plot to show the results of the RSR study between six inflammatory factors and CHD, as shown in Figure 1.

**Table 2.** The RSR of inflammatory factors to CHD

| Inflammatory factors | RSR | Prediction interval |
| --- | --- | --- |
| IL-1β | -13.39 | (-41.71, 12.28) |
| IL-6 | 56.86 | (-192.12, 137.88) |
| TNF-α | 4.97 | (2.990, 10.548) |
| CRP | -14.67 | (-19.27, 240.74) |
| PCT | 21.87 | (2.29, 232.28) |
| SAA | 9.66 | (-85.98, 14.02) |

Note: RSR: risk score ratio; IL-1β : interleukin-1β ; IL-6: interleukin-6; TNF-α : tumor necrosis factor-α ; CRP: C-reactive protein; PCT: procalcitonin; SAA: serum amyloid A.

**Figure 1.**
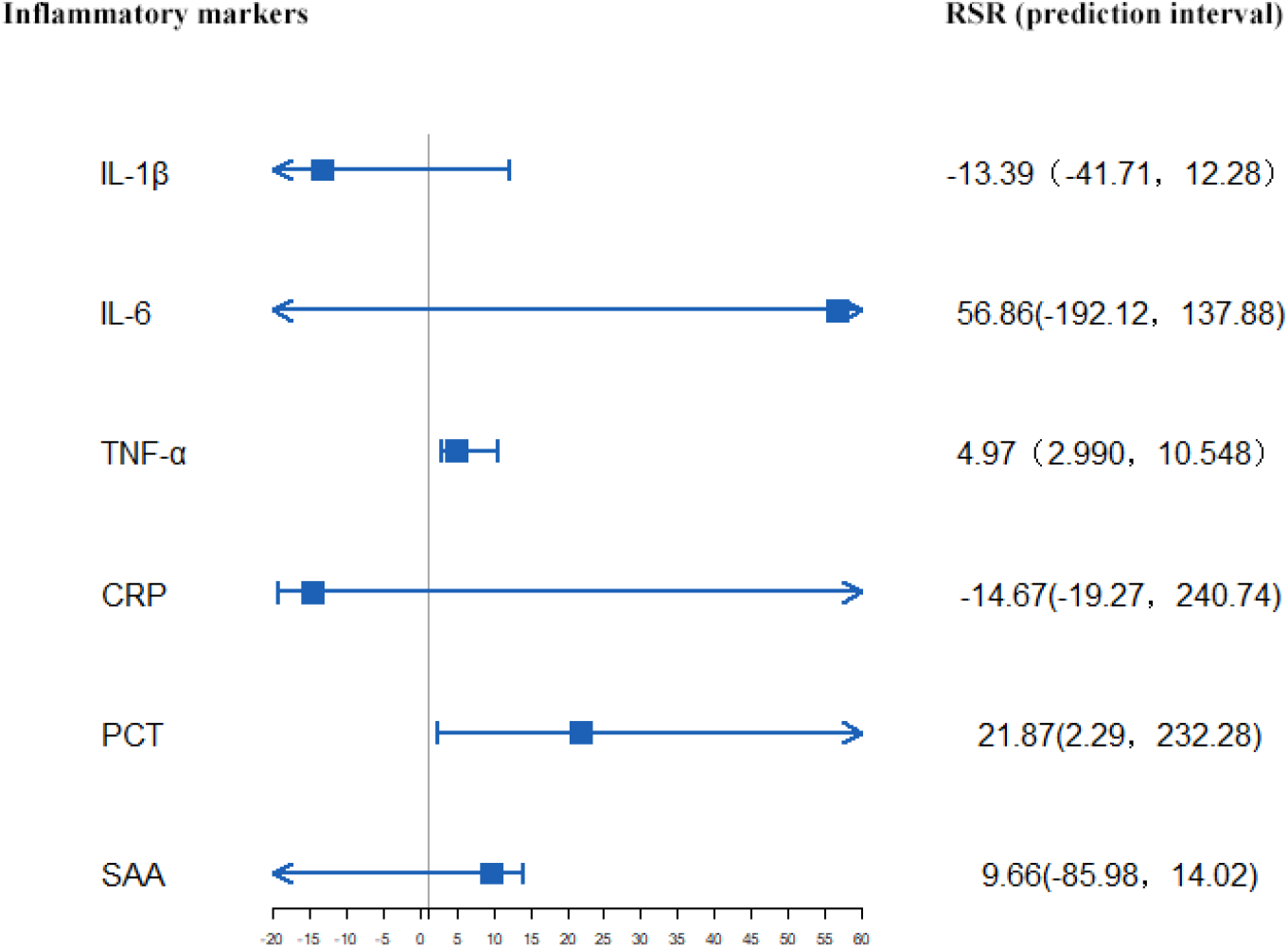
Forest plot of the RSR of inflammatory factors to CHD Note: RSR: risk score ratio; CHD: coronary heart disease; IL-1β: interleukin-1β; IL-6: interleukin-6; TNF-α: tumor necrosis factor-α; CRP: C-reactive protein; PCT: procalcitonin; SAA: serum amyloid A.

## 4 Discussion

Risk prediction and exploration of potential etiology play a crucial role in disease prevention and control. Although the traditional RCT test is the “gold standard” for etiology determination, its practical application is limited by ethics, manpower, financial resources and time. On the other hand, non-randomized studies are increasingly used in practice due to the availability of data and ease of implementation. However, in observational studies, the potential bias of confounders on the accuracy and validity of the results cannot be ignored^[30]^. In this study, starting from the PRS, we extended the disease risk assessment based on genetic information from individuals to groups. Based on population genetic information, we propose PMPRS to assess the risk of disease in a sample population. Further, based on the PMPRS, we propose an RSR for the exploration of potential risk factors for the disease.

With the rapid development of genomics technology, public summary data of gene sequencing related to various diseases will become more easily available, which will provide more convenience for researchers to use genetic information to explore the etiology of diseases. RSR is a research tool for inferring etiological clues based on genetic information. Benefiting from the unique characteristics of genes, that the role of genes in the occurrence and development of diseases is hardly affected by confounding and reverse causality, the power of RSR estimation depends more on the proportion of genes that can explain the disease and the sample size of the original GWAS. However, the majority of known human diseases are affected by both genes and external environmental factors. Moreover, due to the existence of compensatory responses that individuals may initiate during growth and development, the accuracy of RSR results will be affected^[31]^. For the above reasons, we believe that the level of etiological evidence obtained by RSR is higher than that of observational studies, but lower than that of RCT studies. When RCTs cannot be carried out due to limitations of research time, funding, and ethics, RSR can be simply and conveniently used for the initial exploration of disease etiology, and it can providing directions for subsequent research. In addition, while the researcher gives the results of etiology exploration based on traditional observational studies, it is recommended to perform RSR studies at the same time as a mutual verification of different methods in etiology exploration.

CHD is a chronic disease caused by a variety of risk factors. The influence of inflammatory factors in the occurrence and development of CHD has been controversial for a long time. In this study, we divided six common inflammatory markers into “upstream” and “downstream” inflammatory factors, and used PMPRS and RSR to explore their associations with CHD. We found that among the “upstream” inflammatory markers, TNF-α is a potential risk factor for CHD, which is consistent with previous studies^[32]^. Studies have confirmed that endothelial cell damage is the initiating factor of atherosclerosis and CHD, and TNF receptors on vascular endothelial cells provide a direct condition for TNF to damage vascular endothelial cells^[33]^. Moreover, TNF not only damages vascular endothelial cells, but also further damages target cells by activating lysosomes and proteases to promote degranulation of neutrophils^[34]^. Among the “downstream” inflammatory markers, our findings support an association between PCT and CHD. Erren et al reported that elevated PCT concentrations were associated with the degree of atherosclerosis in patients with CHD and peripheral arterial disease^[35]^. Furthermore, the mechanism underlying the relationship between serum PCT and coronary atherosclerotic burden may be related to the increase of inflammatory state^[36]^. Studies have shown that PCT plays an important role in the process of monocyte adhesion and migration, and further stimulates the gene expression of inducible nitric oxide synthase in patients with sepsis, which is a key process of atherosclerotic plaque formation in the occurrence and development of CHD^[36, 37]^. Local or systemic inflammation with continuous monocyte activation is a prerequisite for PCT production^[38]^. As a chemotactic agent, PCT is initially produced in adherent monocytes and subsequently recruits parenchymal cells of inflamed tissue to further produce PCT^[37, 39]^. Peripheral blood mononuclear cells directly stimulated PCT mRNA expression via lipopolysaccharide or indirectly via proinflammatory cytokines IL-1β, IL-6 and TNF-α. The above processes also play an important role in the occurrence and development of CHD^[40]^. However, in this study, although the RSR results showed that the risk score ratio of IL-6 to CHD was as high as 56.86, the prediction interval results did not support the association between IL-6 and CHD. Part of the reason for this result may be that genetic ecidence does not support the association between IL-6 and CHD. It is also possible that the RSR follows the Cauchy distribution and cannot give the expectation and variance. We applied the Boorstrap method to estimate the prediction interval by resamples 1000 times, and this would lose the accuracy of the estimation results.

In this study, based on the polygenic risk score, we proposed the PMPRS to make disease risk prediction from the individual level to the population level, and we also proposed the RSR for the exploration of the potential etiology of the disease. There are following limitations in above methods need to be further studied: First of all, the linkage disequilibrium between genes was not considered in the study, which may bias the accuracy of the estimated results when the genes included in the study have linkage disequilibrium problems. Secondly, since the RSR follows the Cauchy distribution, the variance of the RSR cannot be obtained, and that means the prediction interval of the RSRcannot be estimated directly. The accuracy of the prediction interval estimated by the bootstrap method currently used in this study is small, and the accuracy of the RSR estimation and its interval estimation should be further improved in future research. Finally, in the study of the relationship between CHD and inflammatory factors, this study only provided evidence from the perspective of genetics. In the future, the role of inflammatory factors in the occurrence and development of CHD should be more comprehensively explained from the perspective of external environment and pathogenesis.

## 5 Conclusion

Starting from the PRS, this study constructed the PMPRS for population disease risk assessment, and further developed the RSR for disease etiology exploration. The study used PMPRS and RSR to explore the relationship between CHD and six inflammatory markers in the European population. The results showed that TNF-α and PCT can be considered as potential risk factors in the development of CHD from a genetic point of view. The results of the study have not confirmed the association between IL-1β, IL-6, CRP, SAA and CHD from a genetic point of view.

## Data Availability

All data produced are available online at https://gwas.mrcieu.ac.uk/ and https://www.uk10k.org/

## Acknowledgements

The authors wish to thank all of the investigators for sharing GWAS data. We sincerely acknowledge the UK Biobank, the INTERVAL study and the UK10K study.

## Author’s contributions

SMH conceived the study, performed the statistical analyses, and drafted the manuscript. LQP and JRJ acquired data and helped interpret the data. JPW participated in revising the manuscript. TW was responsible for the concept, design of the study, and critical revisions of the manuscript.

